# Long-Term Prognosis and Early Clinical Predictors in Acute Myocarditis: Insights from a 10-year, Unselected Hospital Cohort

**DOI:** 10.1101/2025.09.08.25335378

**Authors:** Marie Björkenstam, Emanuele Bobbio, Christian L Polte, Niklas Bergh, Michael Fu, Bert Andersson, Clara Hjalmarsson, Entela Bollano

## Abstract

**Background:** Acute myocarditis (AM) is an inflammatory cardiac condition with variable prognosis, ranging from complete recovery to progressive heart failure (HF) or death. While advances in diagnostics, such as high-sensitivity troponins and cardiac magnetic resonance imaging (CMR), have facilitated detection of milder cases, prognostic insights across unselected, real-world populations remain limited.

**Methods:** In this retrospective cohort study, we analyzed 471 consecutive patients hospitalized with a discharge diagnosis of AM between 2009 and 2019. Inclusion was based on clinical diagnosis. Baseline clinical features, laboratory data, and imaging results were extracted and systematically reviewed. The primary composite outcome comprised all-cause mortality, heart transplantation, use of mechanical circulatory support, new-onset HF, ventricular arrhythmias, and cardiac device implantation.

**Results:** The median age was 34 years, 32% were female. Chest pain was the predominant presenting symptom (87%), while ST-segment elevation was observed in 48% of cases. At admission, 24.2% of patients had hypokinesia on echocardiography, and 11.6% had a left ventricular ejection fraction (LVEF) <50%. Within the first year of follow-up, 41 patients (8.7%) experienced the composite outcome. Older age, dyspnea at presentation, and elevated biomarkers were associated with adverse events in the first year. Over a median follow-up of 8.2 years, multivariable analysis revealed age (HR 1.05 per year, 95% CI: 1.03–1.07, p<0.001), signs of HF (HR 3.27, 95% CI: 1.25–8.52, p=0.015), and hypokinesia on echocardiography (HR 19.77, 95% CI: 4.10–95.36, p<0.001) as independent predictors of poor outcomes. No sex-based difference in the primary outcome was observed (HR 0.78, 95% CI: 0.31-1.96, p=0.592) despite different clinical presentation.

**Conclusions:** In this real-world, decade-long cohort, the majority of patients had favorable outcomes. However, a small subset experienced poor outcomes in both the short and long term, highlighting the need for early risk stratification and targeted management of high-risk individuals.

**What is new?:**

- This is one of the largest real-world cohorts (n = 471) of patients hospitalized with acute myocarditis (AM) diagnosed through clinical judgment, CMR, or EMB over a 10-year period.
- Unlike prior studies, inclusion was not limited to CMR-or biopsy-confirmed cases, offering a more representative clinical picture.
- Most patients (87.9%) had favorable long-term outcomes; early adverse events were significantly associated with: older age; heart failure signs at admission and regional hypokinesia.

**What are the clinical implications?:**

- Bedside clinical features, particularly heart failure symptoms and echocardiographic hypokinesia are powerful early predictors of poor prognosis and can guide triage and monitoring decisions.
- Patients without chest pain, or presenting with dyspnea, elevated NT-proBNP, and regional wall motion abnormalities, are at higher risk and may require closer follow-up.
- This study offers a more inclusive and practical understanding of AM prognosis and supports risk-adapted strategies in real-world cardiology.

## Introduction

Acute myocarditis (AM) is an inflammatory condition of the myocardium characterized by a wide spectrum of clinical presentations. Long-term outcomes vary widely, ranging from complete recovery to the development of advanced heart failure (HF), the need for heart transplantation, or death.^1–3^ The severity of the initial inflammatory insult and the extent of myocardial damage largely determine prognosis^2^.

Historically, much of the knowledge regarding AM prognosis has stemmed from biopsy-centric cohorts, typically reflecting individuals with severe disease. As a result, the traditionally reported relatively high mortality rates and risk of HF development may not reflect the full spectrum of patients seen in contemporary clinical practice^4–6^. In real-world settings, the diagnosis of AM is typically based on a combination of clinical presentation and non-invasive diagnostic findings^7–9^. Although endomyocardial biopsy (EMB) remains the gold standard, it is rarely performed in routine practice due to its invasive nature and limited availability^1^ ^10^. The diagnostic landscape has evolved substantially with the introduction of high-sensitivity troponins as well as cardiac magnetic resonance imaging (CMR), allowing for the identification of milder cases that would previously have gone undetected^11–13^. Recent research suggests a benign prognosis in many such milder cases^4^ ^14^ ^15^. However, existing studies often focus on selected populations (e.g., CMR-or EMB-verified AM), and may not reflect the variation seen in general practice, where diagnostic workup varies, and confirmation is inconsistent. Moreover, even among patients with seemingly mild presentations, excess mortality has been reported up to a decade after the initial event,^16^ underscoring the need for improved risk stratification. Current prognostic markers, such as arrhythmias, signs of HF, and left ventricular ejection fraction (LVEF) <50%, are primarily validated in selected cohorts, and their relevance in broader, unselected populations remains unknown^4^ ^14^ ^15^.

In this study, we sought to address this evidence gap by examining a large, unselected cohort hospitalized with clinically suspected AM at a tertiary centre over a 10-year period. By including patients diagnosed through clinical evaluation, CMR, and EMB, we aimed to capture the spectrum of cases encountered in routine practice. Our objectives were to describe clinical features, diagnostic workup, and long-term outcomes, and to identify early prognostic markers that may guide risk stratification and follow-up decisions in this heterogeneous population.

## Methods

### Study Design and Population

This retrospective cohort study included patients from a Swedish university hospital that serves both as the primary referral center for its local catchment area and as a tertiary care center for advanced cases within the region. All consecutive patients > 16 years hospitalized with a primary discharge diagnosis of acute myocarditis (ICD-10 code I40.0, I40.1, I40.8, I40.9, I41.0, I41.1, I41.2, I41.8 or I01.2) between January 1, 2009, and December 31, 2019, were identified. All diagnoses were confirmed through review of medical records to ensure consistency with clinical AM diagnosis. The start of the study period was determined by the publication of the Lake Louise criteria for CMR-based diagnosis of AM in 2009^17^. After exclusion of cases due to alternate diagnoses or loss to follow-up, a final cohort of 471 patients was included in the analysis (**Figure 1**).

**Figure 1.**
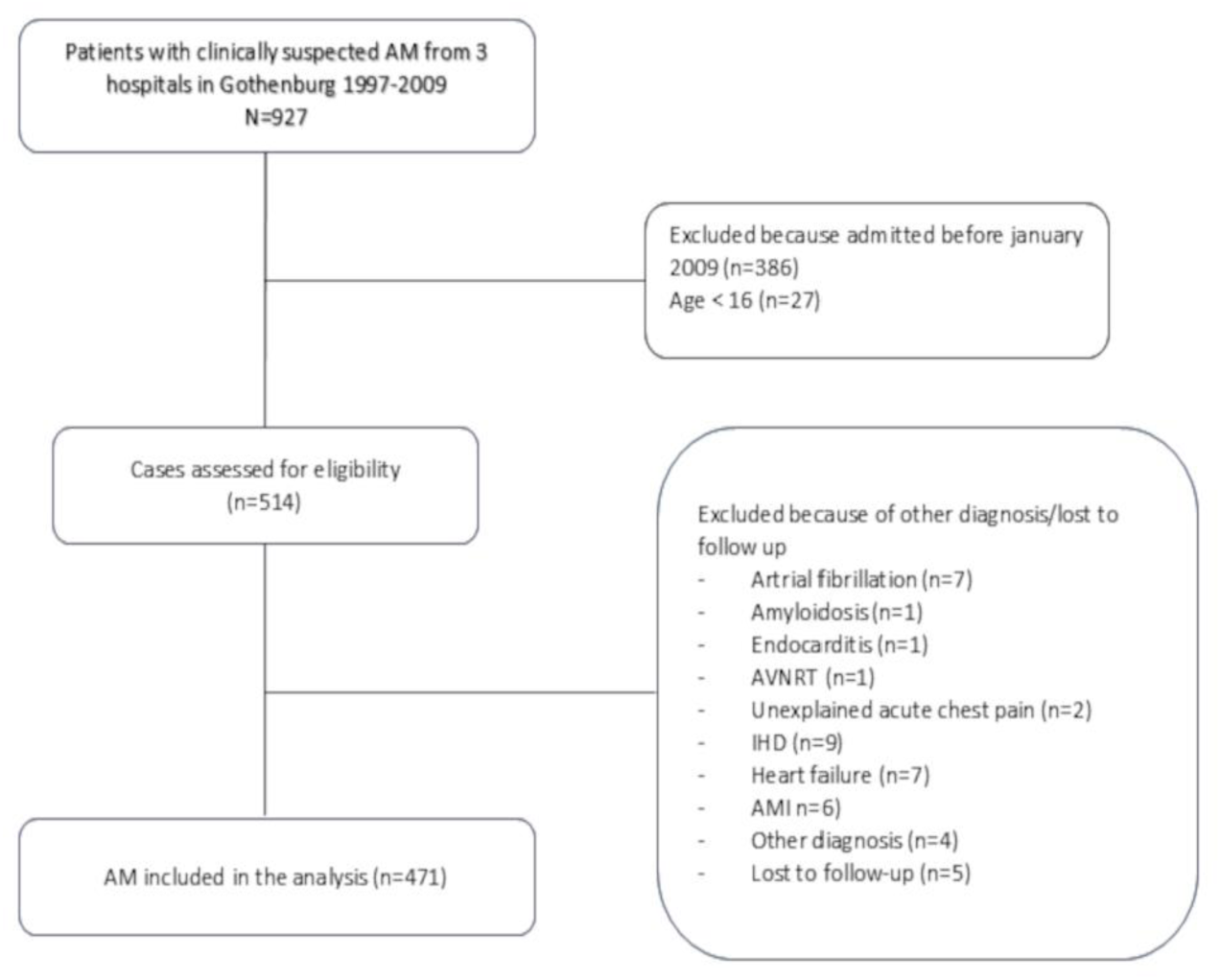
Flow chart illustrating the selection of the study cohort. AM, acute myocarditis; AMI, acute myocardial infarction; AVNRT, AV-nodal re-entry tachycardia; IHD, ischemic heart disease

### Data Collection

Baseline clinical characteristics, including presenting symptoms, physical examination findings, electrocardiogram (ECG), laboratory parameters (e.g., high-sensitivity cardiac troponin [hs-cTn], C-reactive protein [CRP], N-terminal pro–B-type natriuretic peptide [NT-proBNP]), imaging studies (echocardiography and CMR when available) and EMB (which was performed in selected patients) were extracted from electronic medical records.

Echocardiographic assessments were reviewed to determine LVEF, wall motion abnormalities, and presence of pericardial effusion. CMR data were collected regarding whether the examination had been performed, whether findings were consistent with AM and the measured LVEF^13^. Data were collected by trained reviewers. Missing values were not imputed, and cases with missing data were retained in the analyses.

### Outcome Measures

The primary composite outcome included all-cause mortality, orthotopic heart transplantation, use of mechanical circulatory support (MCS), new-onset HF, sustained ventricular arrhythmias, and implantation of an implantable cardioverter-defibrillator (ICD) or pacemaker. Each component of the composite endpoint was treated equally for the purpose of time-to-event analysis. Follow-up data were obtained from national registries and hospital records. Median follow-up was 8.2 years (interquartile range [IQR]: 5.0–11.6 years). Patients were followed until occurrence of an outcome event or end of follow-up (December 2022).

### Statistical Analysis

The results were reported in line with the “Strengthening the Reporting of Observational Studies in Epidemiology” (STROBE) guidelines^18^.

Continuous variables are presented as mean ± standard deviation or median and interquartile range, according to data distribution. Group differences were assessed by Student’s t-test, Mann-Whitney U test, chi-square test, or Fisher’s exact test, as appropriate. Kaplan-Meier survival curves estimated event-free survival, and multivariable Cox proportional hazards regression identified independent predictors of adverse outcomes (candidate variables listed in **Supplementary Table 1**). Variables for multivariate analysis were selected based on clinical relevance and significance in univariate testing. Multivariable Cox regression was performed including time-dependent covariates to account for violations of the proportional hazards assumption, allowing hazard ratios to vary over time. Analyses were performed using IBM SPSS Statistics version 29 (IBM Corp, Armonk, NY, USA) and R version 4.5.0 (R Foundation for Statistical Computing, Vienna, Austria). Statistical significance was set at p<0.05.

### Ethics

The study was approved by the Swedish Ethical Review Authority (Dnr: 026-16) and conformed to the Declaration of Helsinki.

## Results

### Baseline Characteristics

Among 471 included patients, the median age was 34 years (IQR: 23–44), and 32% (n=149) were female. The prevalence of comorbidities was generally low, with 1.9% (n=9) having diabetes, 6.5% (n=30) hypertension, and 3.6% (n=17) a rheumatological disease. A previous diagnosis of heart failure was reported in 3.6% (n=17) of cases. Baseline demographics, clinical presentation, and diagnostic findings are presented in **Table 1**. The most common presenting symptom was chest pain (n=412, 87%), followed by dyspnea (n=88, 20%). A history suggestive of infection within two weeks prior to admission was noted in 65.3% (n=301), and 29.4% (n=125) had fever at admission.

**Table 1.**
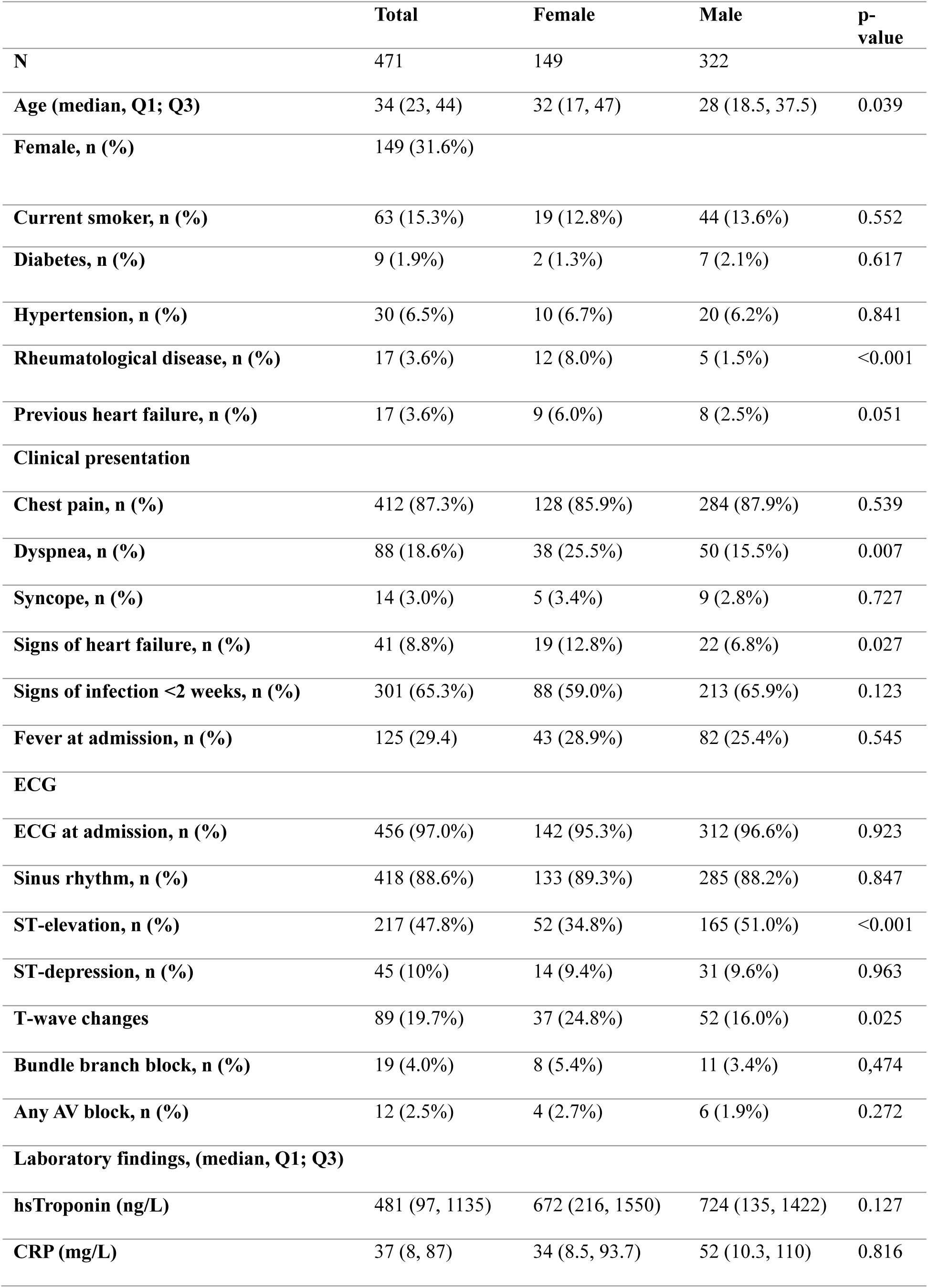
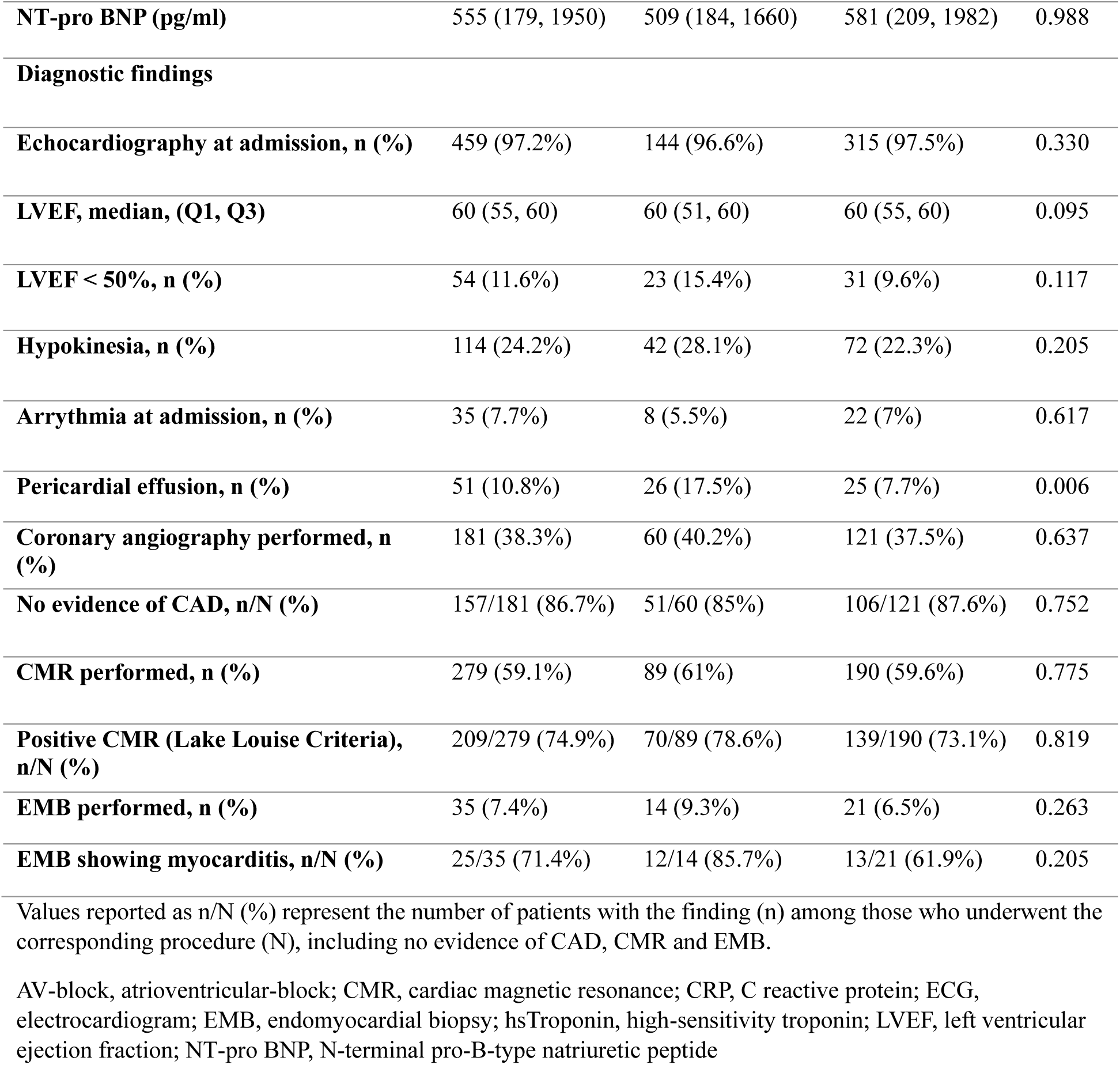
Baseline characteristics, clinical presentation, and diagnostic findings stratified by sex.

On admission ECG, sinus rhythm was observed in 88.6% (n=418) and ST-segment elevation in 47.8%. (n=217). T-wave changes were present in 19.7% (n=89), while bundle branch block and atrioventricular block were infrequent, occurring in 4.0% (n=19) and 2.5% (n= 12), respectively.

Laboratory findings revealed elevated biomarkers: median high-sensitivity troponin was 481 ng/L (IQR: 97–1135), CRP was 37 mg/L (IQR: 8–87), and NT-proBNP was 555 pg/mL (IQR: 179–1950).

Echocardiography was performed in 97.2% (n=459) of patients, with a median LVEF of 60% (IQR: 55–60). Hypokinesia was present in 24.2% (n= 114), and 11.6% (n=54) demonstrated LVEF below 50%. Pericardial effusion was detected in 10.8%, (n=51) and arrhythmia was documented at admission in 7.7% (n=35).

Coronary angiography was performed in 38.3% (n=181) of patients, and in 69.4% (n=59) of those >50 years of age, with no significant coronary artery disease identified in 87.2% of those cases.

Among the *5*9.1% (n=279) of patients who underwent CMR, 75.7% (n=209) showed findings consistent with myocarditis. The use of CMR increased over time, with 33.1% of patients undergoing CMR in the earliest admission quartile (2009-2012) compared to 76,9% in the most recent quartile (2016-2019). EMB was performed in 7.6% (n=36) of patients, yielding histological confirmation of myocarditis in 70.3% (n=25), including eosinophilic myocarditis (n=2), giant cell myocarditis (n=6) and sarcoidosis (n=2).

As summarized in **Table 1**, women were older than men (median age 32 vs 28 years, p<0.039) and more frequently had a history of rheumatological disease (8.0% vs 1.5% p<0.001). Dyspnea (25.5% vs. 15.5%, p=0.007) and signs of heart failure (12.8% vs. 6.8%, p=0.027) were more common in women, while men presented more often with chest pain and ST-segment elevation. T-wave abnormalities were more frequently observed in women (24.8% vs. 16%, p=0.025). Laboratory markers were similar between sexes. Both sexes underwent similar diagnostic work-up. LVEF and regional hypokinesia rates did not differ, but pericardial effusion was more common in women (17.5% vs. 7.7%, p=0.006).

### Short-term Outcomes

Within the first year of follow-up, 41 patients (8.7%) experienced the composite outcome, which included all-cause mortality, heart transplantation, use of mechanical circulatory support, new-onset heart failure, ventricular arrhythmias, or cardiac device implantation (**figure 2**).

**Figure 2.**
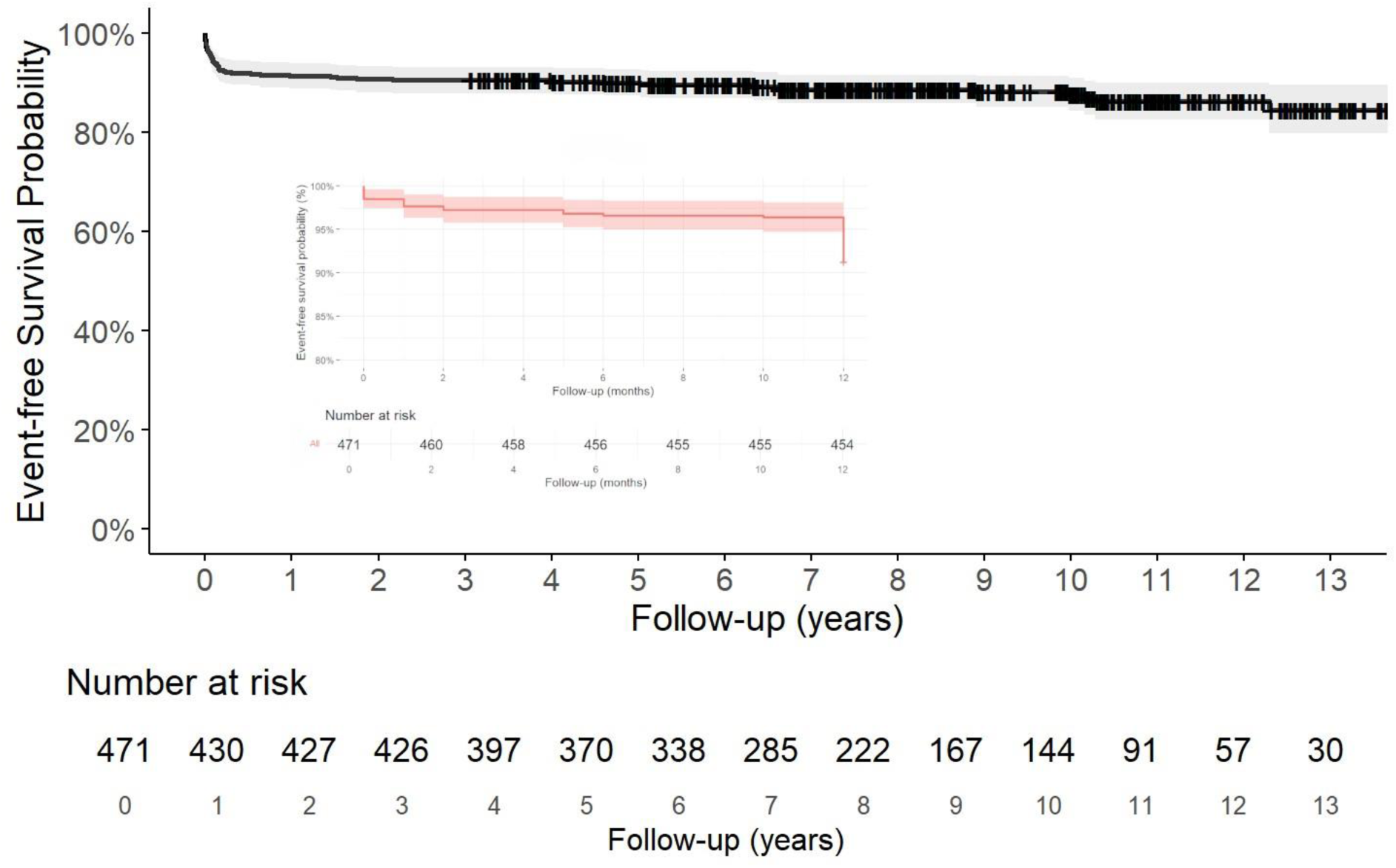
Kaplan-Meier curves illustrating event-free survival for the composite endpoint. The main panel depicts survival over the entire follow-up period. The inset shows event-free survival during the first year, with the y-axis truncated at 80–100% to better visualize differences during this period. Patients were censored at 12 months if event-free.

Patients who experienced an event within the first year were significantly older (median age 52 vs. 28 years, *p*<0.001), and more often had comorbidities such as hypertension (14.6% vs. 5.6%, *p*=0.031) and rheumatological disease (9.7% vs. 3.0%, *p*=0.047). Chest pain was less common among these individuals (46.3% vs. 91.4%; *p*<0.001) while dyspnea (39% vs. 16.7%, *p*<0.001) and clinical signs of heart failure (6.3% vs. 3.3%, *p*<0.001) were markedly more frequent in the event group.

The patients who encountered an outcome also exhibited higher levels of biomarkers (median NT-proBNP 7190 vs. 401 pg/mL, p<0.001; troponin 1071 vs. 672 ng/L, p=0.006; CRP 50 vs. 41 mg/L, p=0.043) and worse echocardiographic findings (median LVEF 30% vs. 60%, p<0.001; LVEF <50% in 73.2% vs. 5.6%, p<0.001; hypokinesia in 73.2% vs. 19.5%, p<0.001; pericardial effusion in 31.7% vs. 8.8%, p<0.001) compared to those without early events (**table 2**).

**Table 2.** Baseline characteristics, clinical presentation and diagnostic findings by event status during the first year of follow-up.

|  | All | Event during first year | No event during first year | p-value |
| --- | --- | --- | --- | --- |
| **N** | 471 | 41 | 430 |  |
| **Age (median; Q1, Q3)** | 34 (23, 44) | 52 (38, 67) | 28 (19, 37) | <0.001 |
| **Female, n (%)** | 149 (31.6%) | 16 (39%) | 133 (30.1%) | 0.287 |
| **Current smoker, n (%)** | 63 (15%) | 7 (17%) | 56 (13%) | 0.418 |
| **Hypertension, n (%)** | 30 (6.5%) | 6 (14.6%) | 24 (5.6%) | 0.031 |
| **Rheumatological disease, n (%)** | 17 (3.6%) | 4 (9.7%) | 13 (3.0%) | 0.047 |
| **Previous heart failure, n (%)** | 17 (3.6%) | 9 (21.9%) | 8 (1.9%) | <0.001 |
| **Clinical presentation** |  |  |  |  |
| **Chest pain, n (%)** | 412 (87.3%) | 19 (46.3%) | 393 (91.4%) | <0.001 |
| **Dyspnea, n (%)** | 88 (18.6%) | 16 (39%) | 72 (16.7%) | <0.001 |
| **Syncope, n (%)** | 14 (3.0%) | 2 (4.9%) | 12 (2.8%) | 0.301 |
| **Signs of heart failure, n (%)** | 40 (8.5%) | 26 (6.3%) | 14 (3.3%) | <0.001 |
| **Signs of infection <2 weeks, n (%)** | 300 (65.7%) | 21 (51.2%) | 279 (64.9%) | 0.260 |
| **ECG** |  |  |  |  |
| **Sinus rhythm, n (%)** | 419 (88.9%) | 26 (63.4%) | 393 (91.4%) | <0.001 |
| **ST-elevation, n (%)** | 217 (47.8%) | 13 (31.7%) | 204 (47.4%) | 0.498 |
| **ST-depression, n (%)** | 45 (10%) | 4 (9.8%) | 41 (9.5%) | 0.570 |
| **T-wave changes** | 89 (19.7%) | 6 (14.6%) | 83 (19.3%) | 0.961 |
| **Bundle branch block, n (%)** | 19 (4.0%) | 3 (7.3%) | 16 (3.7%) | 0.095 |
| **AV-block III, n (%)** | 6 (1.3%) | 2 (4.9%) | 4 (0.9%) | 0.005 |
| **Laboratory findings, (median, Q1; Q3)** |  |  |  |  |
| **hsTroponin (ng/L)** | 719 (75, 1363) | 1071 (187, 1955) | 672 (7, 1337) | 0.006 |
| **CRP (mg/L)** | 42 (19, 65) | 50 (0.5, 100) | 41 (19, 63) | 0.043 |
| **NT-pro BNP (pg/ml)** | 540 (113, 966) | 7190 (4530, 9850) | 401 (141, 660) | <0.001 |
| **Diagnostic findings** |  |  |  |  |
| **Echocardiography at admission, n (%)** | 465 (98.7%) | 39 (95.1%) | 420 (97.7%) | 0.478 |
| **LVEF, median, (Q1, Q3)** | 60 (55, 60) | 30 (17, 43) | 60 (58, 62) | <0.001 |
| **LVEF <50%, n (%)** | 54 (11.6%) | 30 (73.2%) | 24 (5.6%) | <0.001 |
| **Hypokinesia, n (%)** | 114 (24.2%) | 30 (73.2%) | 84 (19.5%) | <0.001 |
| **Pericardial effusion, n (%)** | 51 (10.8%) | 13 (31.7%) | 38 (8.8%) | <0.001 |
| **Coronary angiography performed, n (%)** | 181 (38.3%) | 26 (63.4%) | 155 (36%) | 0.011 |
| **No evidence of CAD, n/N (%)** | 157/181 (86.7%) | 21/26 (80.7%) | 136/155 (87.7%) | 0.324 |
| **CMR performed, n (%)** | 279 (59.1%) | 25 (60.1%) | 254 (59%) | 0.866 |
| **Positive CMR (Lake Louise Criteria), n/N (%)** | 209/279 (74.9%) | 20/25 (80%) | 189/254 (74.4%) | 0.518 |
| **EMB performed, n (%)** | 35 (7.4%) | 26 (63.4%) | 9 (2.1%) | <0.001 |
| **EMB showing myocarditis, n/N (%)** | 25/35 (71.4%) | 20/26 (76.9%) | 5/9 (55.6%) | 0.205 |

Values reported as n/N (%) represent the number of patients with the finding (n) among those who underwent the corresponding procedure (N), including no evidence of CAD, CMR and EMB.
AV-block; atrioventricular-block; CAD, coronary artery disease; CMR, cardiac magnetic resonance; CRP, C reactive protein; ECG, electrocardiogram; EMB, endomyocardial biopsy; hsTroponin, high-sensitivity troponin; LVEF, left ventricular ejection fraction; NT-pro BNP, N-terminal pro-B-type natriuretic peptide

### Long-Term Outcomes

During a median follow-up of 8.2 years (IQR: 5.0–11.6), 57 patients (12.1%) reached the composite outcome (**figure 2**). **Table 3** shows the distribution of the first occurring event per patient.

**Table 3.** Distribution of first events within the composite outcome.

| **Component of the composite outcome**<br>**(first event)** | **N** | **Percentage** | **95%**<br>**Confidence**<br>**interval** |
| --- | --- | --- | --- |
| Cardiac arrest | 4 | 0.8 % | 0.3–2.2 % |
| Death or Heart transplantation | 18 | 3.8 % | 2.4–6 % |
| Incident heart failure | 19 | 4 % | 2.6–6.2 % |
| Device-implantation (ICD/PM) | 6 | 1.3 % | 0.6–2.8 % |
| Mechanical circulatory support<br>(LVAD/ECMO) | 7 | 1.5 % | 0.7–3 % |
| Ventricular tachycardia | 3 | 0.6 % | 0.2–1.9 % |

Each patient is included only once, based on the first occurring event. Percentages refer to the total cohort (n=471). ECMO, extracorporeal mechanical oxygenation; ICD, implantable cardiac defibrillator; LVAD, left ventricular assist device; PM, pacemaker.

In multivariable Cox regression analysis, older age (HR 1.05 per year, 95% CI: 1.03–1.07, p<0.001), signs of heart failure at presentation (HR 3.27, 95% CI: 1.25–8.52, p=0.015), and hypokinesia on echocardiography (HR 19.77, 95% CI: 4.10–95.36, p<0.001) were independent predictors of adverse events (**table 4**). No significant sex-based difference in the composite outcome was observed (HR 0.78; 95% CI 0.31–1.96, p=0.592).

Although reduced LVEF (<50%) and the absence of chest pain were associated with adverse outcomes in univariate analysis, these did not remain significant in the adjusted model.

Notably, sex, ST-elevation, and and the performance of a CMR examination (analysed as a binary variable independent of imaging findings) did not independently predict outcomes.

Kaplan–Meier survival analysis revealed distinct separation of event-free survival curves based on presence of hypokinesia and HF signs at baseline, emphasizing the prognostic relevance of early bedside echocardiographic assessment (**Figure 3**).

**Figure 3.**
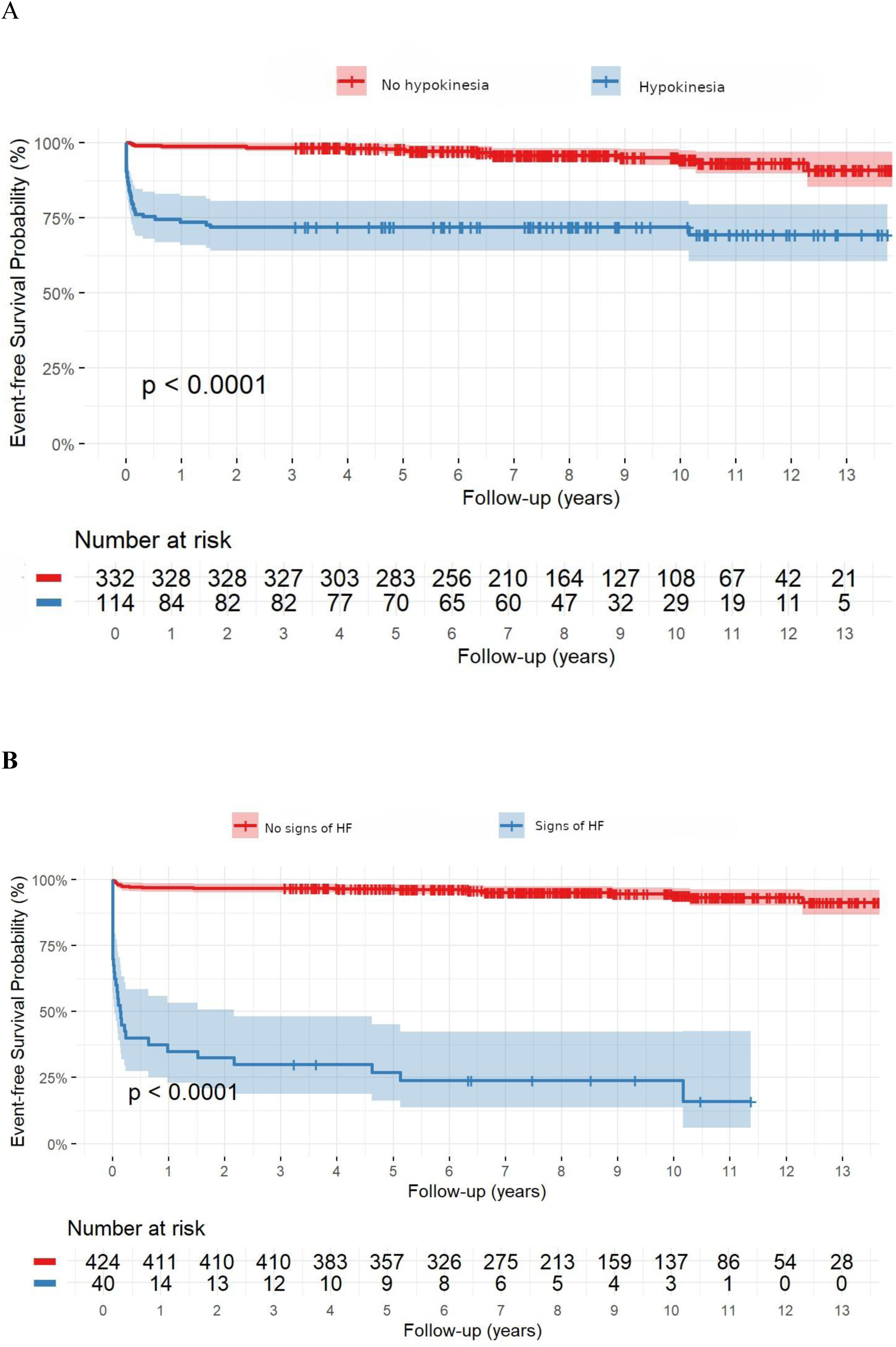
Kaplan–Meier curves for event-free survival stratified by **(A)** baseline hypokinesia and **(B)** heart failure (HF) signs.

## Discussion

In this comparatively large retrospective cohort study spanning over a decade, we evaluated long-term outcomes of 471 patients hospitalized with AM at a university hospital. Unlike previous studies, the cohort was not limited to CMR-or EMB-confirmed cases. By including all clinically diagnosed patients, we captured a broader and more representative range of disease presentations, reflective of everyday practice.

Our data demonstrate that most individuals with AM experience favorable long-term prognosis, with 87.9% remaining free from all-cause mortality, incident HF, heart transplantation, ventricular arrhythmias, or device implantation over a median follow-up of 8.2 years. Our analysis revealed three variables that predicted poor outcomes: increasing age, clinical signs of heart failure, and hypokinesia on echocardiography. All of which are easily assessable early during hospitalization, reinforcing the prognostic utility of basic bedside and imaging evaluation. Conversely, sex and ECG features such as ST-elevation were not associated with prognosis after adjustment.

### Prognosis and event rates

Our finding that most patients with AM experience favorable long-term outcomes is consistent with observations from CMR-derived studies using the Lake Louise Criteria, which have reported 5-year survival rates above 90% in milder cases^4^ ^11^ ^19^. However, those studies often excluded more severe or unstable patients who could not undergo CMR. On the other hand, EMB-based series report worse outcomes, with survival as low as 56% at 4.3 years^20^. Our real-world approach means that we included severe cases as well as patients otherwise excluded from EMB or imaging-based studies, and thus present a more balanced epidemiologic picture.

### Predictors of adverse outcomes

Older age and clinical signs of HF at admission were strong independent predictors of poor prognosis, echoing observations from prior large-scale cohorts^5^ ^14^. Ammirati et al. reported similar associations, emphasizing the prognostic weight of hemodynamic compromise and ventricular dysfunction at presentation^4^. Our finding that regional hypokinesia was a particularly powerful predictor (HR 19.77) adds valuable granularity. While global LVEF has traditionally been emphasized, focal wall motion abnormalities may better reflect early or localized myocardial involvement, especially in patients with near-normal LVEF^9^ ^21^.

Elevated NT-proBNP and troponin concentrations were also linked to short-term adverse outcomes, further supporting their utility in risk stratification, in line with existing literature,^4^ ^14^ ^15^ ^22^ ^23^. However, in adjusted analyses, hypokinesia and signs of HF remained the strongest predictors for adverse outcomes. Interestingly, LVEF <50%, a commonly used threshold, did not independently predict long-term outcomes in our cohort, possibly due to high cut-off value or variability in echocardiographic assessment. LVEF is a crude marker of systolic function and lacks sensitivity to detect myocardial inflammation, which may be better reflected by hypokinesia, which, thus, may be a better predictor of prognosis^1^ ^17^.

### Chest pain and clinical phenotypes

The presenting symptom of chest pain was inversely associated with the composite outcome, as seen in previous studies. ^4^ ^14^ ^23^ ^24^ This presentation may reflect a subepicardial or pericardial inflammatory process rather than transmural damage, suggesting a more favorable course^25^ ^26^. Conversely, patients presenting with dyspnea, elevated natriuretic peptides, and reduced LVEF likely exhibit a more fulminant or diffuse myocardial involvement. Interestingly, only 46% of patients who experienced the composite outcome during the first year presented with chest pain, compared to 91% of those without events, highlighting the prognostic importance of atypical presentations.

### Sex Differences: Phenotype vs. Prognosis

Consistent with the literature, our cohort showed a male predominance (68.4%)^2^ ^27^. Men more often presented with chest pain and ST-elevation, whereas women more frequently had dyspnea, pericardial effusion, and a higher prevalence of rheumatological disease, possibly reflecting a greater contribution of autoimmune mechanisms^28^. Despite these phenotypic differences^14^ ^29–31^, sex was not associated with the long-term risk of experiencing the composite outcome, reinforcing the idea that sex may shape the clinical phenotype more than the outcome in AM^14^. This contrasts with other studies that have shown that female gender is related to a high risk for myocarditis-related complications and in-hospital mortality^32^.

### Comparison with more highly selected cohorts

Previous studies limited to CMR-confirmed AM report favorable outcomes but may underestimate event rates by excluding unstable patients who cannot tolerate imaging^8^ ^19^ ^33^. CMR was performed in 59% of patients (33.1% in the first quartile vs 76.9% in the last), reflecting increasing availability over the study period. In our cohort, undergoing CMR *per se* was not associated with outcomes. This likely reflects selection bias: patients in stable condition were more likely to undergo CMR, while those with fulminant disease commonly required urgent management without advanced imaging. It should be emphasized that our analyses did not account for imaging findings, but rather focused on whether or not a CMR examination was performed. Therefore, these results should not be interpreted as evidence regarding the prognostic value of CMR-derived parameters.

Conversely, EMB-based cohorts may overestimate risk, since biopsy is often reserved for fulminant or refractory cases^8^ ^10^. In summary, our study bridges the gap between highly selected, imaging-based cohorts and the diverse, real-world population encountered in general cardiology.

### Clinical implications and future directions

These results have direct clinical relevance as they suggest that bedside assessment, particularly for HF signs and hypokinesia, remains essential in AM. Patients presenting with dyspnea or elevated biomarkers, especially in the absence of chest pain, should prompt heightened vigilance and closer monitoring.

Prospective validation in larger cohorts is needed, especially to define follow-up strategies and to assess whether repeat imaging or serial biomarkers add prognostic value.

### Strengths and Limitations

This study’s strengths include its large, unselected real-world cohort, consecutive patient inclusion over a decade, and extended follow-up period with few patients lost to follow-up. This enhances the reliability and generalizability of our findings, particularly compared to highly selected CMR-or EMB-based studies. However, limitations inherent to its retrospective design exist. The fact that not all cases were verified by CMR or EMB, may have led to potential diagnostic misclassification, although previous work has shown high coding validity for myocarditis in Swedish hospital records (>80%)^34^. Some adverse events outside our institution could have been missed, although national registry linkage mitigated this risk. Despite these limitations, our results are broadly consistent with contemporary AM cohorts and contribute important real-world insight.

## Conclusions

In this unselected, real-world cohort, AM was associated with generally favourable long-term outcomes. However, older age, signs of HF at presentation, and regional hypokinesia were independently associated with poor prognosis. These findings emphasize the importance of timely bedside evaluation, especially in settings with limited access to advanced imaging.

Overall, these findings support a pragmatic, inclusive approach to risk assessment and management in patients admitted with AM.

## Non-standard Abbreviations and Acronyms

AM: Acute myocarditis
CMR: Cardiac magnetic resonance
CRP: C-reactive protein
EMB: Endomyocardial biopsy
HF: Heart failure
hs-cTn: High-sensitivity cardiac troponin
ICD: Implantable cardioverter-defibrillator
IQR: Interquartile range
LVEF: Left ventricular ejection fraction
MCS: Mechanical circulatory support
NT-proBNP: N-terminal pro–B-type natriuretic peptide
STROBE: Strengthening the Reporting of Observational Studies in Epidemiology

## Data Availability

The datasets generated and/or analyzed during the current study are available from the corresponding author on reasonable request

## Acknowledgments

The authors have no acknowledgments.

---

**Supplementary Table 1.** Selection of variables for multivariable analysis based on univariate analysis.

| **Variable** | **Univariate**<br>**HR (95% CI)** | **p-value** | **Multivariate HR**<br>**(95% CI)** | **p-value** |
| --- | --- | --- | --- | --- |
| **Age** | 1.05; 1.04-1.06 | <0.001 | 1.05 (1.03 – 1.07) | <0.001 |
| **Female sex** | 0.8; 0.46-1.39 | 0.425 | - |  |
| **History of HF** | 11.2; 5.86-21.30 | <0.001 |  |  |
| **Hypertension** | 2.7; 1.29-5.79 | 0.009 |  |  |
| **Diabetes** | 1.9; 0.74-4.83 | 0.180 | - |  |
| **Rheumatological**<br>**disease** | 4.82; 2.17-10.7 | <0.001 |  |  |
| **AT ADMISSION** |  |  |  |  |
| **Chest pain** | 0.13; 0.07-0.23 | <0.001 | 0.99 (0.41 – 2.31) | 0.974 |
| **Dyspnea** | 2.68; 1.52-4.7 | <0.001 |  |  |
| **Signs of HF** | 29.98; 17.1-52.6 | <0.001 | 3.27 (1.25 – 8.52) | 0.015 |
| **ST-elevation** | 0.72; 0.4-1.3 | 0.267 | - |  |
| **T-wave changes** | 1.09; 0.54-2.2 | 0.812 | - |  |
| **Troponin** | 1.22; 1.0-1.47 | 0.045 |  |  |
| **LVEF < 50%** | 124 (56-275) | <0.001 | 0.95 (0.28-3.23) | 0.940 |
| **Hypokinesia** | 7.25; 3.99-13.18 | <0.001 | 19.77 (4.10-95.36) | <0.001 |
| **Pericardial effusion** | 3.16; 1.59-6.24 | <0.001 | 0.93 (0.30-2.89) | 0.992 |
| **Positive CMR**<br>**(Lake Louise**<br>**Criteria)** | 1.41; 0.75-2.65 | 0.288 |  |  |

The table presents results from univariate analyses assessing the association between baseline characteristics, clinical variables, and outcomes. A maximum of 6 variables were included in the multivariate Cox regression due to the number of adverse events in line with statistical guidelines for model stability
CMR, cardiac magnetic resonance; HF, heart failure; LVEF, left ventricular ejection fraction

